# Risk factors for retirement home COVID-19 outbreaks in Ontario, Canada: A population-level cohort study

**DOI:** 10.1101/2020.12.08.20246124

**Authors:** Andrew P. Costa, Derek R. Manis, Aaron Jones, Nathan M. Stall, Kevin A. Brown, Veronique Boscart, Adriane Castellino, George A. Heckman, Michael P. Hillmer, Chloe Ma, Paul Pham, Saad Rais, Samir K Sinha, Jeffrey W. Poss

## Abstract

**Background:** The epidemiology of COVID-19 in retirement homes (also known as assisted living facilities) is largely unknown. We examined the association between retirement home and community level characteristics and the risk of COVID-19 outbreaks in retirement homes during the first wave of the COVID-19 epidemic.

**Methods:** We conducted a population-based retrospective cohort study of licensed retirement homes in Ontario, Canada, from March 1st - September 24th, 2020. Our primary outcome was a COVID-19 outbreak (≥1 resident or staff confirmed case by validated nucleic acid amplification assay). We used time-dependent proportional hazards methods to model the associations between retirement home and community level characteristics and COVID-19 outbreaks.

**Results:** Our cohort included all 770 licensed retirement homes in Ontario, which housed 56,491 residents. There were 172 (22.3%) COVID-19 retirement home outbreaks involving 1,045 (1.9%) residents and 548 staff (1.5%). COVID-19 cases were distributed unevenly across retirement homes, with 1,593 (92.2%) resident and staff cases occurring in 77 (10%) of homes. The adjusted hazard of a COVID-19 outbreak in a retirement home was positively associated with homes that had a large resident capacity, homes that were co-located with a long-term care facility, large corporate owned chains, homes that offered many services onsite, increases in regional COVID-19 incidence, and a higher community-level ethnic concentration.

**Interpretation:** Readily identifiable retirement home-level characteristics are independently associated with COVID-19 outbreaks and may support risk identification. A higher ethnic concentration of the community surrounding a retirement home is associated COVID-19 outbreaks, with an uncertain mechanism.

## Introduction

Frail older adults living in congregate care settings have been at the centre of the Coronavirus Disease 2019 (COVID-19) pandemic in Canada and internationally (1,2). Long-term care (LTC) facilities—the congregate living population most affected by COVID-19—have been the subject of immense scientific and public interest during the pandemic (3). Retirement homes (often known as assisted living facilities) have received far less examination despite also housing a large population of vulnerable older adults (4–7). Retirement homes are predominantly private residential complexes that provide a range of services (e.g., ADL assistance, meals, medication management) that are typically less-intensive compared to LTC facilities. They are almost exclusively financed through out-of-pocket payments by residents or their families (8), and the lack of consistent regulation throughout Canada and the United States has limited research into the epidemiology of COVID-19 in retirement homes (9).

There are close to 800 licensed retirement homes in Canada’s most populous province of Ontario that house over 50,000 older adults, a population size that approaches the number of Ontario LTC home residents (10). Since the onset of the COVID-19 pandemic in Ontario, the number of positive cases and deaths in retirement homes continued to grow. As of November 16th, 2020, retirement home residents accounted for approximately 8% of COVID-19 deaths in Ontario (287/3505) (11). COVID-19 outbreaks have surged in retirement homes during the second wave in Canada and the United States (11,12), with limited examination beyond early reports of case identification (13).

We examined the association between home and community level characteristics and the risk of COVID-19 outbreaks during the first wave of the COVID-19 epidemic in Ontario’s retirement homes. Consistent with our previous population-level work in Ontario LTC homes (2,14), we hypothesized that home size and regional COVID-19 incidence would be associated with the risk of an outbreak.

## Methods

### Study design

We conducted a retrospective cohort study across all retirement homes in Ontario, Canada from March 1st, 2020 until September 24^th^ 2020; spanning the entirety of Ontario’s first wave of the COVID-19 pandemic (15). In Ontario, retirement homes are defined in legislation (similar to other North American jurisdictions) as residential complexes that are occupied primarily by persons who are 65 years of age or older and have at least two of thirteen care services available (Appendix 1)(16,17). Our study was approved by the Hamilton Integrated Research Ethics Board. We adhered to the Strengthening the Reporting of Observational Studies in Epidemiology (STROBE) reporting guideline and the Reporting of Studies Conducted Using Observational Routinely-Collected Health Data (RECORD) statement guidelines (Appendix 2) (18,19).

### Data sources

Data used for this study were obtained from the Ontario Retirement Homes Regulatory Authority (RHRA) and Ontario Ministry of Health as part of the COVID-19 Ontario Consensus Modelling Table. We obtained home-level daily COVID-19 case counts and deaths among retirement home residents and staff (in home or hospital) from the RHRA through their COVID Tracking Tool. These data, which include the date on which a COVID-19 outbreak was declared and deemed resolved, were collected through daily direct inquires by the RHRA and through self-reports by licensed retirement homes to the RHRA and have recently been publicly reported (11). Licensed retirement homes are required to report COVID-19 outbreaks to the RHRA at the same time they are reported to their local public health units. Data from the tracking tool correlated closely with other provincial data sources including the integrated Public Health Information System, the Ontario Laboratory Information System, and a death database maintained by the Ontario Chief Coroner’s office, and have been used in COVID-19 research in LTC homes (2,14).

### Exposures

We obtained home-level exposure data from the provincial registry of licensed retirement homes, which contains data on resident capacity, co-location with a LTC facility, and the availability of care services onsite for all retirement homes in Ontario. The RHRA is legislatively mandated to maintain the registry as the provincial retirement homes regulator. Home size was based on reported resident capacity and assigned to quintiles. Co-location of the retirement home with a LTC facility was identified by RHRA records as those homes sharing the same physical building or situated on the same site. Information on chain ownership was supplied by the RHRA, and homes were classified as being members of a small (2 to 5 homes), medium (6 to 20 homes) or large (>20 homes) chain, or not part of a chain. The RHRA maintains a list of 13 services offered (Appendix 1) and a home’s services were summed and assigned to the following categories: ≤6, 7, 8, or ≥9 services.

We obtained data on home level occupancy, staffing counts, and external care providers from an RHRA survey of all retirement homes conducted in May 2020 (home level response = 92.7%). External care providers come into the home either as contracted workers of the publicly funded home care program, or by private pay arrangement with residents. A staff to resident ratio comprising both types of external care providers was calculated based on the home survey response values and assigned to quartiles.

We obtained the daily incidence of COVID-19 across Ontario’s 36 public health regions from Public Health Ontario’s integrated Public Health Information System (20,21). Time varying COVID-19 incidence was calculated each day as the rolling 14-day incidence per 1000 population for the public health unit in which the retirement home resided. We chose a rolling 14-day incidence based on the trend of community incidence rates (See Appendix 3 and 4). We conducted a sensitivity analysis using a 30-day time period, as well as by centering the index day in the rolling average period. We obtained linked data on community-level median household income and ethnic concentration from the Ontario Ministry of Health based on the 2016 Canadian Census and the Ontario Marginalization Index, respectively. We obtained the neighbourhood-level ethnic concentration surrounding each retirement home, which is defined as the combined proportions non-white and non-Indigenous residents and immigrants that arrived in Canada within the past five years, based on the 2016 Canadian Census (22,23). The population size situating each retirement home was calculated using Statistics Canada’s Postal Code Conversion File Plus (PCCF+), using postal codes from the Canada Post Corporation which were current up to and including November 2018; communities with a population size of <10,000 individuals are rural (24).

### Outcomes

Our primary outcome was a COVID-19 outbreak (≥1 resident or staff case confirmed by validated nucleic acid amplification assay)

### Statistical Analysis

Summary statistics were computed to compare, by COVID-19 outbreak status, retirement home and community level characteristics. The Chi-square test was used for categorical variables and the Kruskal-Wallis test was used for continuous variables. Case fatality rates were calculated as the proportion of residents who died of COVID-19 compared to the total number of residents infected with COVID-19.

We used Cox proportional hazards to model the associations between retirement home and community level characteristics and the risk of a COVID-19 outbreak. Community incidence of COVID-19 was a time-varying covariate, with all other measures being fixed. A retirement home was at risk of experiencing an event on all days (March 1 to September 24, 2020), except for days in which the home was experiencing an outbreak.When an outbreak was over the home returned to being at risk for a future outbreak. To account for correlation between observations within public health units and multiple outbreaks within the same retirement homes, we applied a robust sandwich estimator for the covariance matrix. We built multivariable models using a manual, forward selection approach, with community size forced into the model given its strong association with risk of outbreak in previous work. Selection proceeded in ascending order of bivariate p-values, retaining only variables with a p <0.05 in the multivariable model. After the initial forward selection, all unselected variables were again tested individually in the model, and retained if p <0.05. Finally, two-way interactions among selected covariates were examined. We examined the proportionality of hazards assumption using Kaplan-Meier plots and time-varying covariates. Some explanatory variables represented as quintiles were collapsed to three-levels to comply with proportionality requirements.

## Results

### COVID-19 Cases and Deaths in Retirement Homes

The analysis included all 770 licensed retirement homes in Ontario as of March 1st 2020. Overall, 172 (22.3%) retirement homes experienced one or more COVID-19 outbreaks as of September 24^th^ 2020, with 154 (20.0%) having one outbreak and 18 (2.33%) having two outbreaks. COVID-19 outbreaks in this period involved 1,045 (1.9%) infected residents and 548 infected staff (1.5%) (Table 1). Over 90% of outbreaks occurred before June 2020 (Appendix 3). Almost half of all outbreaks involved both staff and resident cases, with outbreaks involving staff cases being more common than those that involved resident cases. The crude cumulative incidence of COVID-19 among residents was 18.5 per thousand. In homes with a resident infection, the median number of residents infected was 2 (IQR: 1-13). COVID-19 infection was distributed unevenly across retirement homes: 1,593 (92.2%) resident and staff cases occurred in 77 (10%) of homes. There were 51 (6.6%) retirement homes with outbreaks resulting in one or more resident deaths, accounting for a total of 215 resident COVID-19 deaths (3.8 per 1,000 residents in Ontario) and a case fatality rate of 20.5%.

**Table 1:** COVID-19 outbreaks and deaths in Retirement Homes, Ontario (Mar. 1-Sep. 24, 2020; N= 770)

|  | All licensed retirement homes (n = 770) |
| --- | --- |
| **COVID-19 Outbreaks N (% facilities)** |  |
| 1+ Outbreaks | 172 (22.3%) |
| Outbreaks involving both residents and staff | 78 (10.1%) |
| Outbreaks involving residents only | 39 (5.1%) |
| Outbreaks involving staff only | 55 (7.1%) |
| **COVID-19 Cases** |  |
| Staff Infections, N | 548 |
| Resident Infections, N | 1,045 |
| Cumulative incidence of COVID-19 resident cases (per thousand RH residents) | 18.5 |
| Proportion of residents infected per home with a resident infection, median (IQR) | 3.0% (1.3% - 13.6%) |
| Median number of cases per home with a resident infection, (IQR) | 2 (1 – 13) |
| **COVID-19 Deaths** |  |
| Homes with 1+ COVID-19 deaths N | 51 |
| COVID-19 deaths N | 215 |
| COVID-19 death rate (per thousand RH residents) | 3.81 |
| Proportion of residents who died per home with a resident death, median (IQR) | 4.2% (1.4% - 8.2%) |
| Median number of deaths per home with one or more deaths (IQR) | 3 (1 – 6) |
| Case fatality rate % | 20.5% |

### Retirement Home Characteristics by Outbreak Status

Retirement homes housed 56,491 residents with an average of approximately 73 residents and 48 staff per home and an overall occupancy rate of 74.2% (Table 2). The majority of retirement homes had a capacity of over 100 residents, had external care providers that entered the home daily, were corporate-owned chains, were located in communities with larger populations and lower ethnic concentration, and offered more than 6 services to their residents on-site. The minority of retirement homes were co-located with a LTC facility. Compared to retirement homes without outbreaks, those that experienced one or more COVID-19 outbreaks were more likely to have larger capacity, be part of a corporate-owned chain, have external care providers entering the home on a daily basis, have more services available to their residents onsite, and be located in larger communities with a higher ethnic concentration.

**Table 2:**
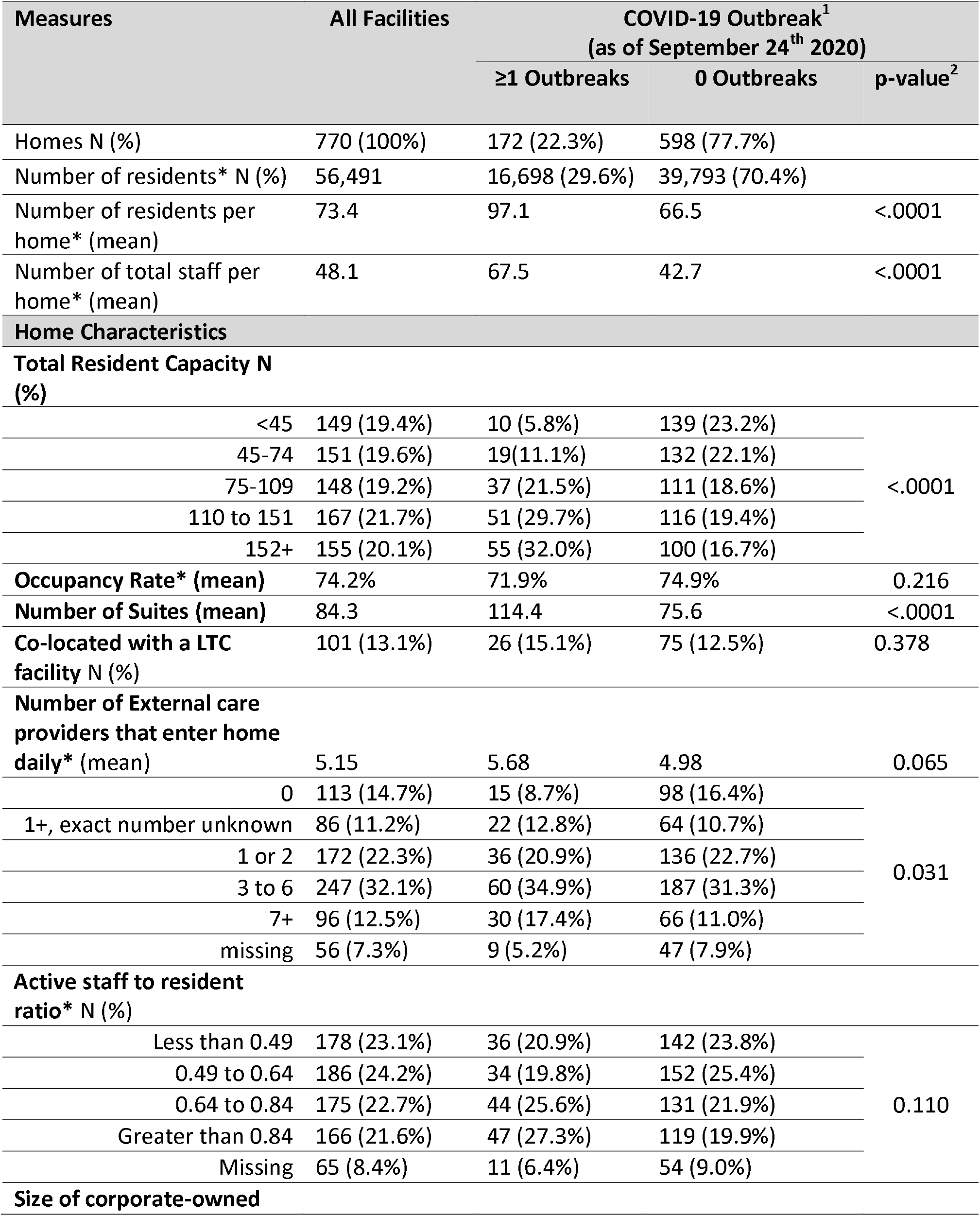
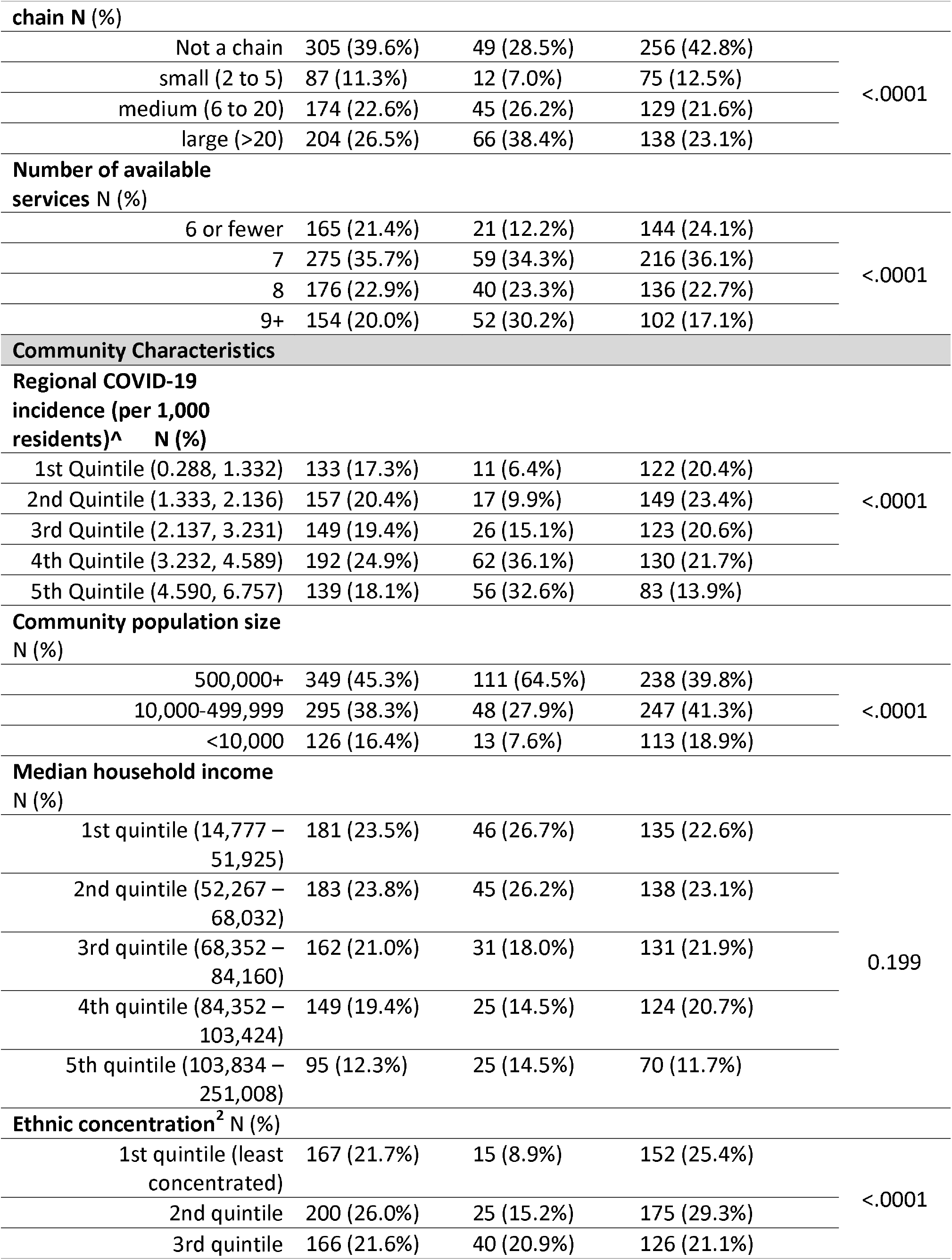
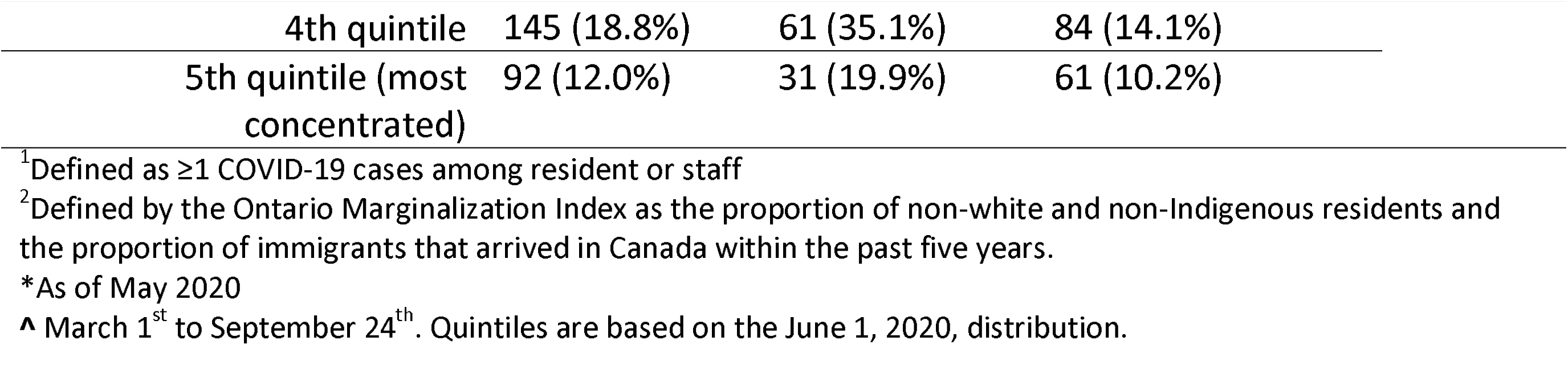
Characteristics of retirement homes by COVID-19 outbreak status, Ontario (Mar. 1-Sep. 24, 2020; N= 770)

### Risk of a COVID-19 outbreak

Increases in regional COVID-19 incidence during the Wave 1 period were strongly associated with COVID-19 outbreaks in retirement homes, where a 1 case per 1,000 person increase in the previous 14 days was associated with a 5.83-fold increase in the hazard of an outbreak (Table 3). In addition to regional COVID-19 incidence, the adjusted hazard of a COVID-19 outbreak was positively associated with homes that had a large resident capacity, large corporate owned chains, homes that were co-located with a LTC facility (adjusted hazard ratio [aHR], 1.84; 95% confidence interval [CI] 1.23-2.74), homes that offered many services onsite, and a higher community-level ethnic concentration. There was pattern of more COVID-19 outbreaks in retirement homes with a higher community-level ethnic concentration across public health regions (Figure 1).

**Table 3.**
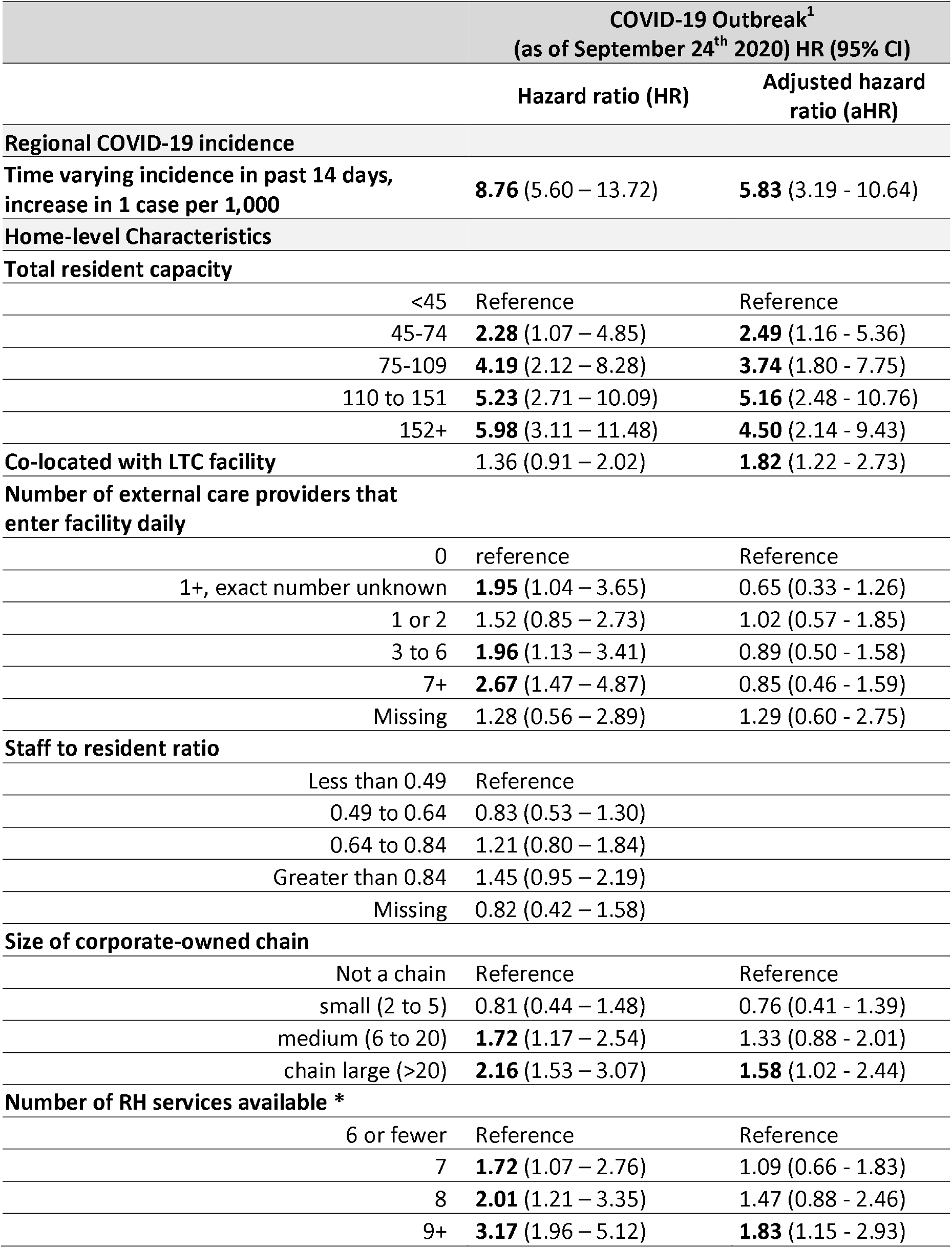
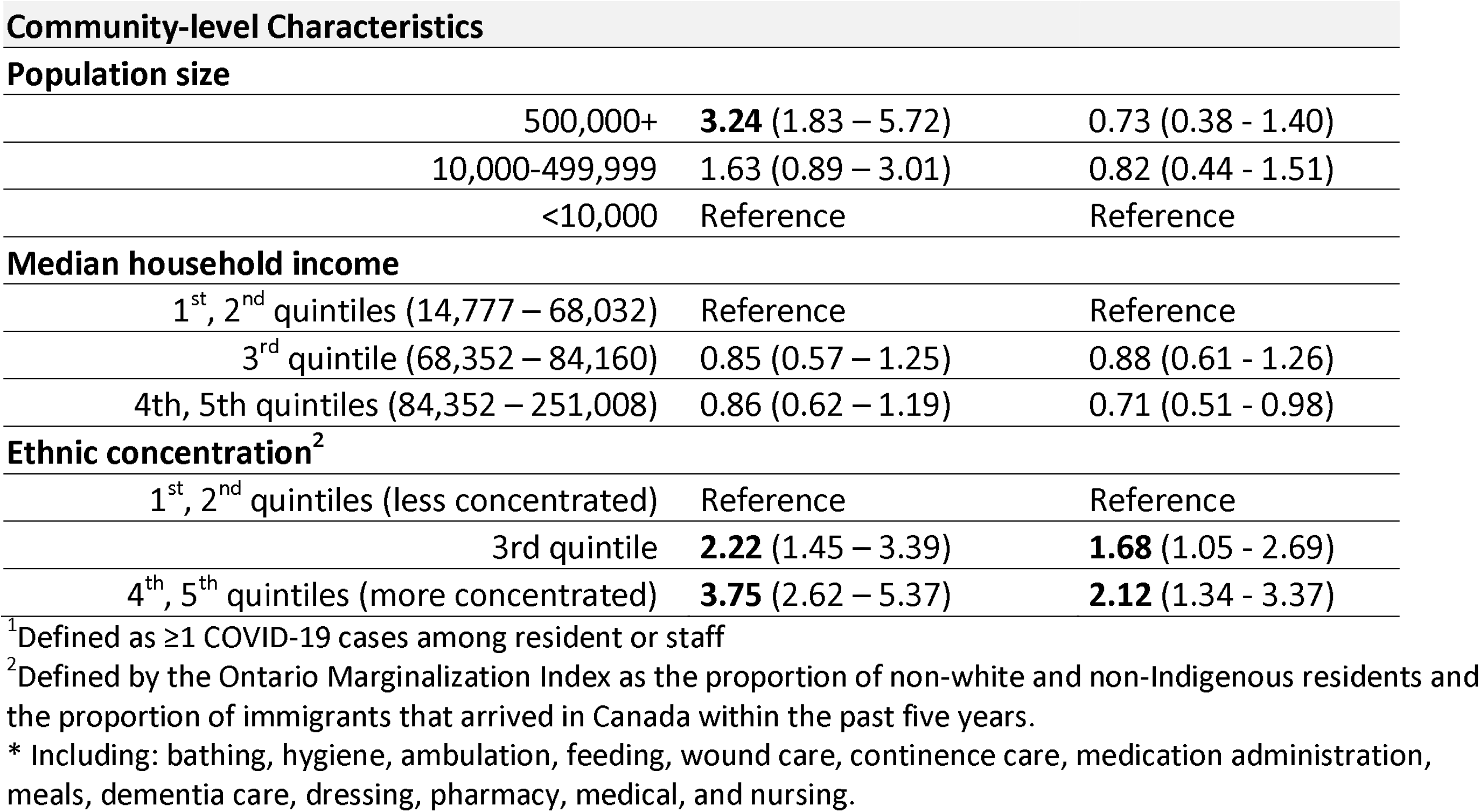
Associations between home and regional characteristics with time to COVID-19 resident outbreak, cumulative COVID-19 cases count, and COVID-19 resident deaths, Ontario, Mar 1-Sep. 24 2020 (N=770 facilities)

**Figure 1.**
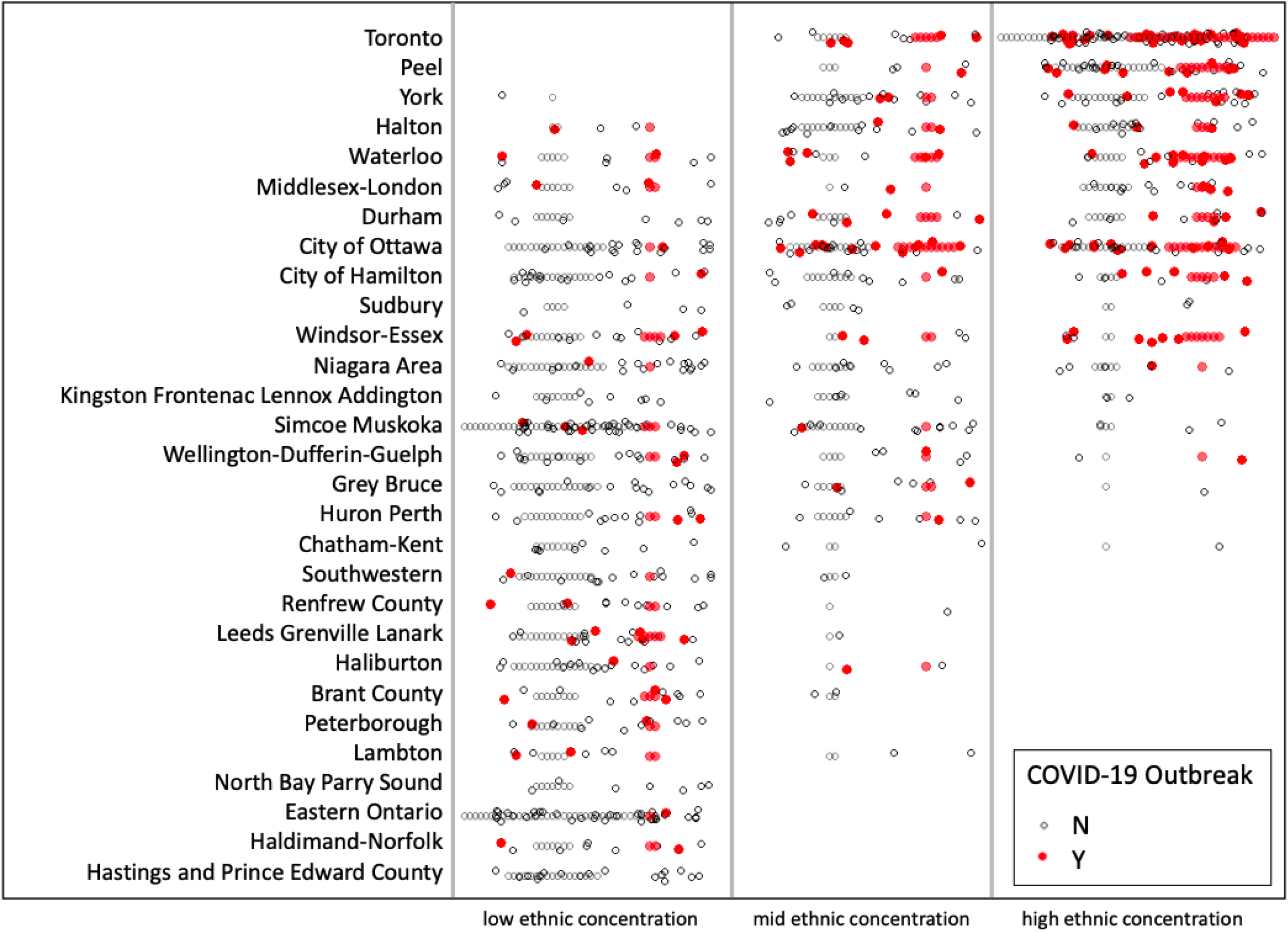
Retirement Home COVID-19 Outbreaks by Community-level Ethnic Concentration and Public Health Unit Region, Ontario, Mar 1-Sep. 24 2020. Public health unit regions with less than 5 retirement homes are excluded.

## Interpretation

In this study of all 770 retirement homes in Ontario, Canada, we found that the hazard of a COVID-19 outbreak was positively associated with increases in regional COVID-19 incidence, homes that had a large resident capacity, homes that were co-located with a LTC facility, large corporate owned chains, homes that offered many services onsite, and a higher community-level ethnic concentration. We identify risk factors for COVID-19 outbreaks in retirement homes that can inform risk identification at the provincial and regional level, as has been done for the LTC sector (25).

Consistent with LTC facilities (26,27), our findings support that the incidence of COVID-19 in a public health region, large chains, and the size of retirement homes are important risk factors for COVID-19 outbreaks. We found a strong association between retirement home outbreaks and the 14-day rolling incidence of COVID-19 in the surrounding public health region, which aligns with the temporal relationship between community COVID-19 incidence and LTC outbreaks (28). Also consistent with the literature on COVID-19 and LTC (2,14), we found that home capacity was associated with risk of COVID-19 outbreaks such that retirement homes with a capacity greater than 100 residents had more than a 5-fold increase in risk of outbreak. Larger homes require more staff, which likely increases the number of potential individuals who are unknowingly importing COVID-19 into homes. Given that the majority of COVID-19 cases were accumulated early in Wave 1 during the period when visitors to retirement homes were severely restricted (Appendix 3), it is probable that retirement home staff and care providers were the main vectors for seeding COVID-19 infection into retirement homes (29). The lack of an adjusted association between the external care providers and COVID-19 outbreak might be explained by the lack of variability between retirement homes of the same size. Homes that offered 9 or more services had an almost 2-fold increase in risk for a COVID-19 outbreak. This likely reflects the additional exposure to SARS-COV-2 associated with additional and more lengthy interactions between staff and residents who require such services.

Retirement homes that are co-located with a LTC facility had an almost 2-fold increase in risk for a COVID-19 outbreak. This finding could reflect staff working between homes, which has been linked to the transmission of COVID-19 between LTC facilities (25,30). Despite provincial orders restricting work at multiple healthcare settings within a 14-day period, emerging evidence suggests that residual connectivity between congregate living settings exists and could represent temporary agency workers and contract staff which are exempted from the provincial order (31). A reduction in mobility of staff between retirement and LTC homes may reduce the risk of COVID-19 outbreaks in co-located retirement homes during successive waves of the COVID-19 pandemic.

We observed an association between levels of ethnic concentration in the community surrounding a retirement home and the risk of COVID-19 outbreak, even after adjusting for regional COVID-19 rates and community level household income. Public Health Ontario reports demonstrate that ethno-culturally dense neighbourhoods experience disproportionately higher rates of COVID-19 (32). LTC facilities that care for more racial and ethnic minority residents report higher COVID-19 cases and/or deaths (33). Our finding may relate, at least partly, to cultural patterns of care and connection for older family members. However, we could not examine the complex relationship between COVID-19 and ethno-cultural characteristics in our analyses, and further data collection and analysis are needed to understand this mechanism.

Comparable to many emerging and rapidly collected sources of COVID-19 data, we could not independently validate completeness with respect to COVID-19 infections and deaths at the home level. We also could not account for temporal changes in infection prevention and control practices and changing provincial policies that may have influenced these results. Our study was limited by the lack of individual level data on clinical, organizational, and sociocultural characteristics that may differ across homes. A better understanding of the complex relationships between COVID-19 and ethno-cultural characteristics require further data at the individual level. Our adjustment of regional COVID-19 incidence may have caused over-adjustment given that some community cases may have been secondary to retirement home cases.

In conclusion, we find that the risk of a COVID-19 outbreak in retirement homes is associated with larger resident capacity, co-location with a LTC facility, large corporate owned chains, a higher availability of services onsite, and increases in regional COVID-19 incidence. For retirement homes co-located with a LTC facility, a reduction in staff mobility between settings is a potentially modifiable factor that may reduce the risk of future COVID-19 outbreaks. Increased ethnic concentration of the community surrounding a retirement home is associated COVID-19 outbreaks, with an uncertain mechanism. Identifying and understanding observed differences in COVID-19 outbreaks across retirement homes may inform risk identification and prevention.

## Data Availability

The study protocol and statistical code are available on request, with the understanding that the computer programs may rely on coding templates or macros that may be inaccessible or require modification. The data sets from this study are held securely at the Retirement Homes Regulatory Authority and Ontario Ministry of Healths Capacity Planning and Analytics Division, and data-sharing agreements prohibit making the data set publicly available.

## Acknowledgements

We gratefully acknowledge the support of Dr. Kamil Malikov of the Ontario Ministry of Health’s Capacity Planning and Analytics Division for assistance with data acquisition. Andrew P. Costa holds the Schlegel Chair in Clinical Epidemiology and Aging at McMaster University. Nathan M. Stall is supported by the Department of Medicine’s Eliot Phillipson Clinician-Scientist Training Program and the Clinician Investigator Program at the University of Toronto, and the Vanier Canada Graduate Scholarship.

---

**Appendix 1:** 13 Care Services Available to Residents (Directly or Indirectly)

| Services |
| --- |
| Assistance with personal hygiene |
| Assistance with bathing |
| Assistance with dressing |
| Assistance with ambulation |
| Assistance with feeding |
| Meal provision |
| Medication administration |
| Dementia care program |
| Continence care |
| Skin and wound care program |
| Nursing services* |
| Medical services^ |
| Pharmacy services~ |

\*Any service that a member of the College of Nurses of Ontario provides while engaging in the practice of nursing.
^Any service that a member of the College of Physicians and Surgeons of Ontario provides while engaging in the practice of medicine.
~Any service that a member of the Ontario College of Pharmacists provides while engaging in the practice of pharmacy.

**Appendix 2:** The RECORD statement – checklist of items, extended from the STROBE statement, that should be reported in observational studies using routinely collected health data

|  | Item No. | STROBE items | Location in manuscript where items are reported | RECORD items | Location in manuscript where items are reported |
| --- | --- | --- | --- | --- | --- |
| **Title and abstract** |  |  |  |  |  |
|  | 1 | (a) Indicate the study's design with a commonly used term in the title or the abstract (b) Provide in the abstract an informative and balanced summary of what was done and what was found | 1 (a) Included, abstract (methods)<br>1 (b) Included, abstract (methods, results, conclusion) | RECORD 1.1: The type of data used should be specified in the title or abstract. When possible, the name of the databases used should be included.<br>RECORD 1.2: If applicable, the geographic region and timeframe within which the study took place should be reported in the title or abstract.<br>RECORD 1.3: If linkage between databases was conducted for the study, this should be clearly stated in the title or abstract. | 1.1 Included, abstract (methods)<br>1.2 Included, title and abstract (methods, results)<br>1.3 N/A |
| **Introduction** |  |  |  |  |  |
| Background rationale | 2 | Explain the scientific background and rationale for the investigation being reported | Included, introduction |  |  |
| Objectives | 3 | State specific objectives, including any prespecified hypotheses | Included, introduction |  |  |
| **Methods** |  |  |  |  |  |
| Study Design | 4 | Present key elements of study design early in the paper | Included, Methods |  |  |
| Setting | 5 | Describe the setting, locations, and relevant dates, including periods of recruitment, exposure, follow-up, and data collection | Included, Methods |  |  |
| Participants | 6 | *(a) Cohort study* - Give the eligibility criteria, and the sources and methods of selection of participants. Describe methods of follow-up<br>*Case-control study* - Give the eligibility criteria, and the sources and methods of case ascertainment and control selection. Give the rationale for the choice of cases and controls<br>*Cross-sectional study* - Give the eligibility criteria, and the sources and methods of selection of participants<br>*(b) Cohort study* - For | 6 (a) Included, Methods | RECORD 6.1: The methods of study population selection (such as codes or algorithms used to identify subjects) should be listed in detail. If this is not possible, an explanation should be provided.<br>RECORD 6.2: Any validation studies of the codes or algorithms used to select the population should be referenced. If validation was conducted for this study and not published elsewhere, detailed methods and results should be provided.<br>RECORD 6.3: If the study involved linkage of databases, consider use of a flow diagram or other | 6.1 Included, Methods<br>6.2 Not Included, data source was not validated – this limitation was highlighted in the discussion<br>6.3 N/A, no data linkages |
|  |  | matched studies, give matching criteria and number of exposed and unexposed<br>*Case-control study* - For matched studies, give matching criteria and the number of controls per case |  | graphical display to demonstrate the data linkage process, including the number of individuals with linked data at each stage. |  |
| Variables | 7 | Clearly define all outcomes, exposures, predictors, potential confounders, and effect modifiers. Give diagnostic criteria, if applicable. | Included, Methods | RECORD 7.1: A complete list of codes and algorithms used to classify exposures, outcomes, confounders, and effect modifiers should be provided. If these cannot be reported, an explanation should be provided. | N/A, no codes or algorithms needed for ascertainment of exposure, outcomes, confounders, and effect modifiers |
| Data sources/<br>measurement | 8 | For each variable of interest, give sources of data and details of methods of assessment (measurement). Describe comparability of assessment methods if there is more than one group | Included, Methods |  |  |
| Bias | 9 | Describe any efforts to address potential sources of bias | Included, Methods |  |  |
| Study size | 10 | Explain how the study size was arrived at | Included, Methods |  |  |
| Quantitative variables | 11 | Explain how quantitative variables were handled in the analyses. If applicable, describe which groupings were chosen, and why | Included, Methods |  |  |
| Statistical methods | 12 | (a) Describe all statistical methods, including those used to control for confounding<br>(b) Describe any methods used to examine subgroups and interactions<br>(c) Explain how missing data were addressed<br>(d) *Cohort study* - If applicable, explain how loss to follow-up was addressed<br>*Case-control study* - If applicable, explain how matching of cases and controls was addressed<br>*Cross-sectional study* - If applicable, describe analytical methods taking account of sampling strategy<br>(e) Describe any sensitivity analyses | 12 (a) Included, Methods (statistical analysis)<br>12 (b) N/A, no subgroups or interactions examined<br>12 (c) No missing data.<br>12 (d) N/A (no loss to follow-up)<br>12 (e) None performed. |  |  |
| Data access and |  | .. |  | RECORD 12.1: Authors should | 12.1 Included, |
| cleaning methods |  |  |  | describe the extent to which the investigators had access to the database population used to create the study population.<br>RECORD 12.2: Authors should provide information on the data cleaning methods used in the study. | Methods (Provincial Modelling Table)<br>12.2 N/A, no data cleaning performed |
| Linkage |  | .. |  | RECORD 12.3: State whether the study included person-level, institutional-level, or other data linkage across two or more databases. The methods of linkage and methods of linkage quality evaluation should be provided. | Included, Methods (institutional-level data used and there were no linkages) |
| **Results** |  |  |  |  |  |
| Participants | 13 | (a) Report the numbers of individuals at each stage of the study (*e.g.*, numbers potentially eligible, examined for eligibility, confirmed eligible, included in the study, completing follow-up, and analysed)<br>(b) Give reasons for non-participation at each stage.<br>(c) Consider use of a flow diagram | 13 (a-c) N/A, all Ontario RH homes included with no loss to follow-up | RECORD 13.1: Describe in detail the selection of the persons included in the study (*i.e.*, study population selection) including filtering based on data quality, data availability and linkage. The selection of included persons can be described in the text and/or by means of the study flow diagram. | N/A, no selection of persons in the study |
| Descriptive data | 14 | (a) Give characteristics of study participants (*e.g.*, | 14 (a) N/A, no RH resident |  |  |
|  |  | demographic, clinical, social) and information on exposures and potential confounders<br>(b) Indicate the number of participants with missing data for each variable of interest<br>(c) *Cohort study* - summarise follow-up time (e.g., average and total amount) | characteristics available (we described home characteristics)<br>14 (b) No missing data<br>14 (c) Included, methods |  |  |
| Outcome data | 15 | *Cohort study* - Report numbers of outcome events or summary measures over time<br>*Case-control study* - Report numbers in each exposure category, or summary measures of exposure<br>*Cross-sectional study* - Report numbers of outcome events or summary measures | Included, Results |  |  |
| Main results | 16 | (a) Give unadjusted estimates and, if applicable, confounder-adjusted estimates and their precision (e.g., 95% confidence interval). Make clear which | 16 (a) Included, Results and Tables |  |  |
|  |  | confounders were adjusted for and why they were included<br>(b) Report category boundaries when continuous variables were categorized<br>(c) If relevant, consider translating estimates of relative risk into absolute risk for a meaningful time period | 16 (b) Included, Results and Tables<br>16 (c) Included, Results and Tables |  |  |
| Other analyses | 17 | Report other analyses done—e.g., analyses of subgroups and interactions, and sensitivity analyses | N/A (no other analyses) |  |  |
| **Discussion** |  |  |  |  |  |
| Key results | 18 | Summarise key results with reference to study objectives | Included, Discussion |  |  |
| Limitations | 19 | Discuss limitations of the study, taking into account sources of potential bias or imprecision. Discuss both direction and magnitude of any potential bias | Included, Discussion | RECORD 19.1: Discuss the implications of using data that were not created or collected to answer the specific research question(s). Include discussion of misclassification bias, unmeasured confounding, missing data, and changing eligibility over time, as they pertain to the study being reported. | Included, Discussion (paragraph on limitations) |
| Interpretation | 20 | Give a cautious overall | Included, |  |  |
|  |  | interpretation of results considering objectives, limitations, multiplicity of analyses, results from similar studies, and other relevant evidence | Conclusion |  |  |
| Generalisability | 21 | Discuss the generalisability (external validity) of the study results | Included, Discussion |  |  |
| **Other Information** |  |  |  |  |  |
| Funding | 22 | Give the source of funding and the role of the funders for the present study and, if applicable, for the original study on which the present article is based | N/A, study was not funded. Funding of individual researcher detailed in the Acknowledgements section |  |  |
| Accessibility of protocol, raw data, and programming code |  | .. |  | RECORD 22.1: Authors should provide information on how to access any supplemental information such as the study protocol, raw data, or programming code. | Included, Acknowledgements Section |

**Appendix 3:**
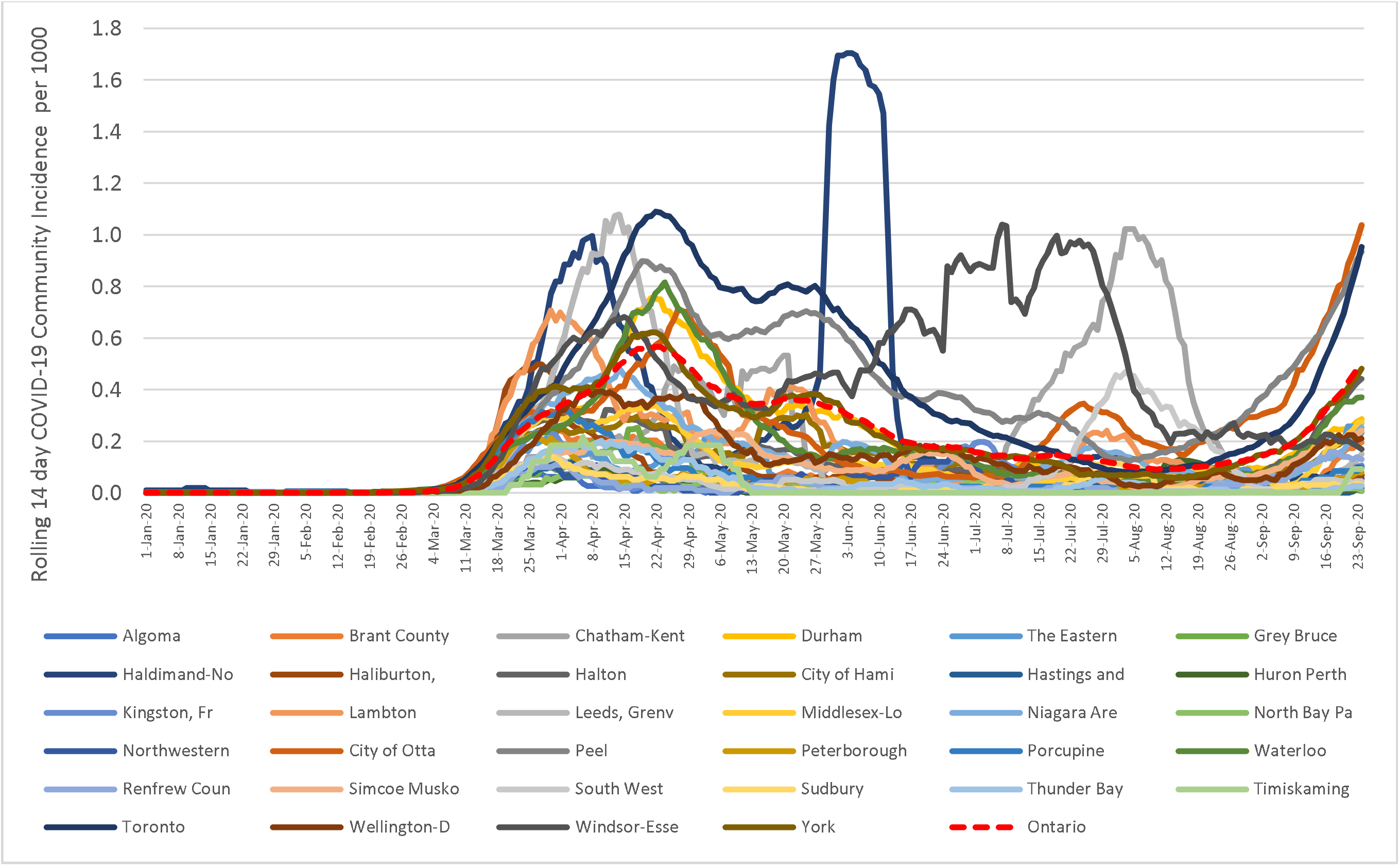
Rolling 14-day COVID-10 case count, by Public Health Unit Region, Ontario, Mar 1-Sep. 24 2020

**Appendix 4:**
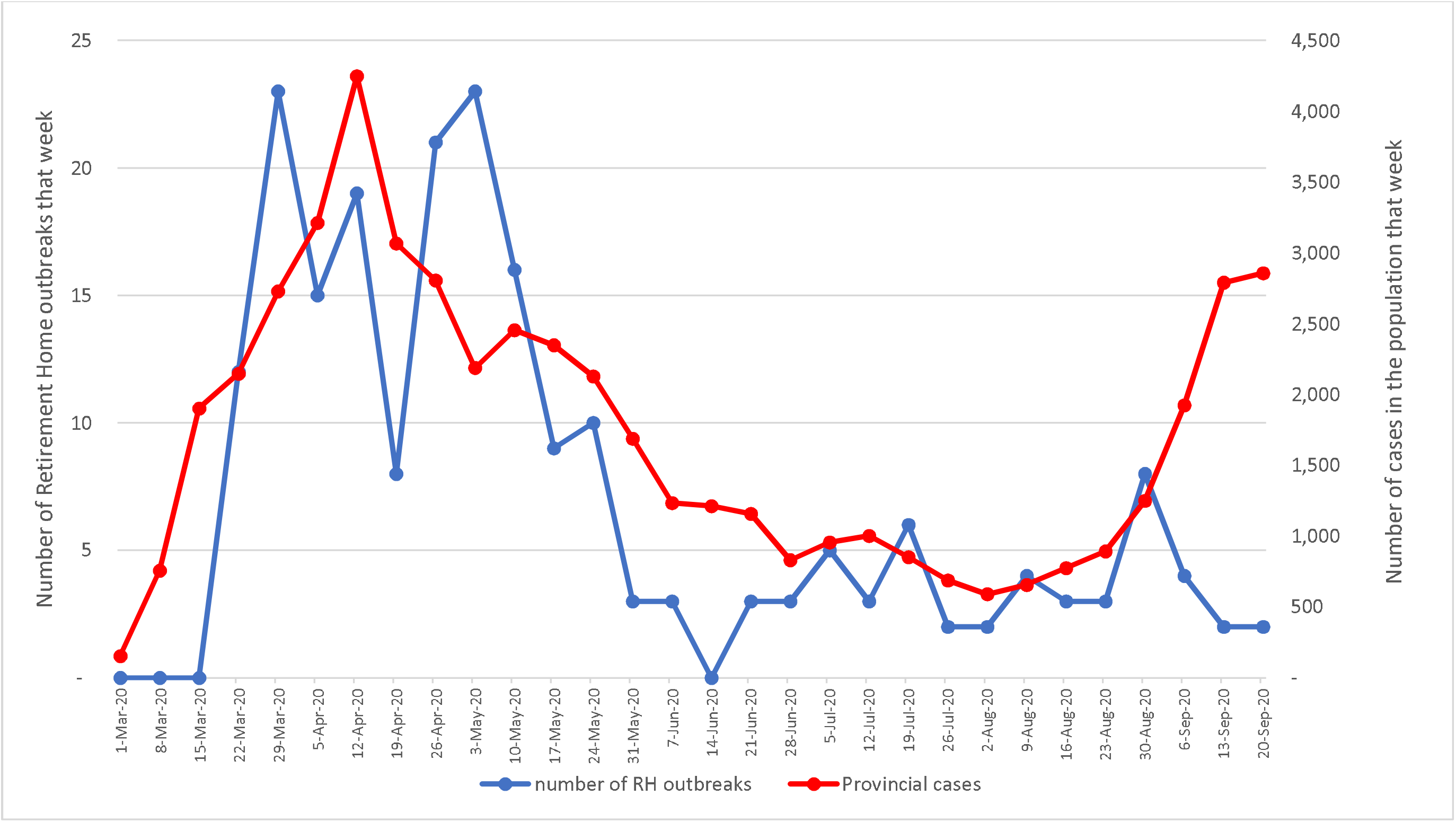
Number of Retirement Home Outbreaks per week, Ontario, Mar 1-Sep. 24 2020 (N=770 facilities)

**Appendix 5:**
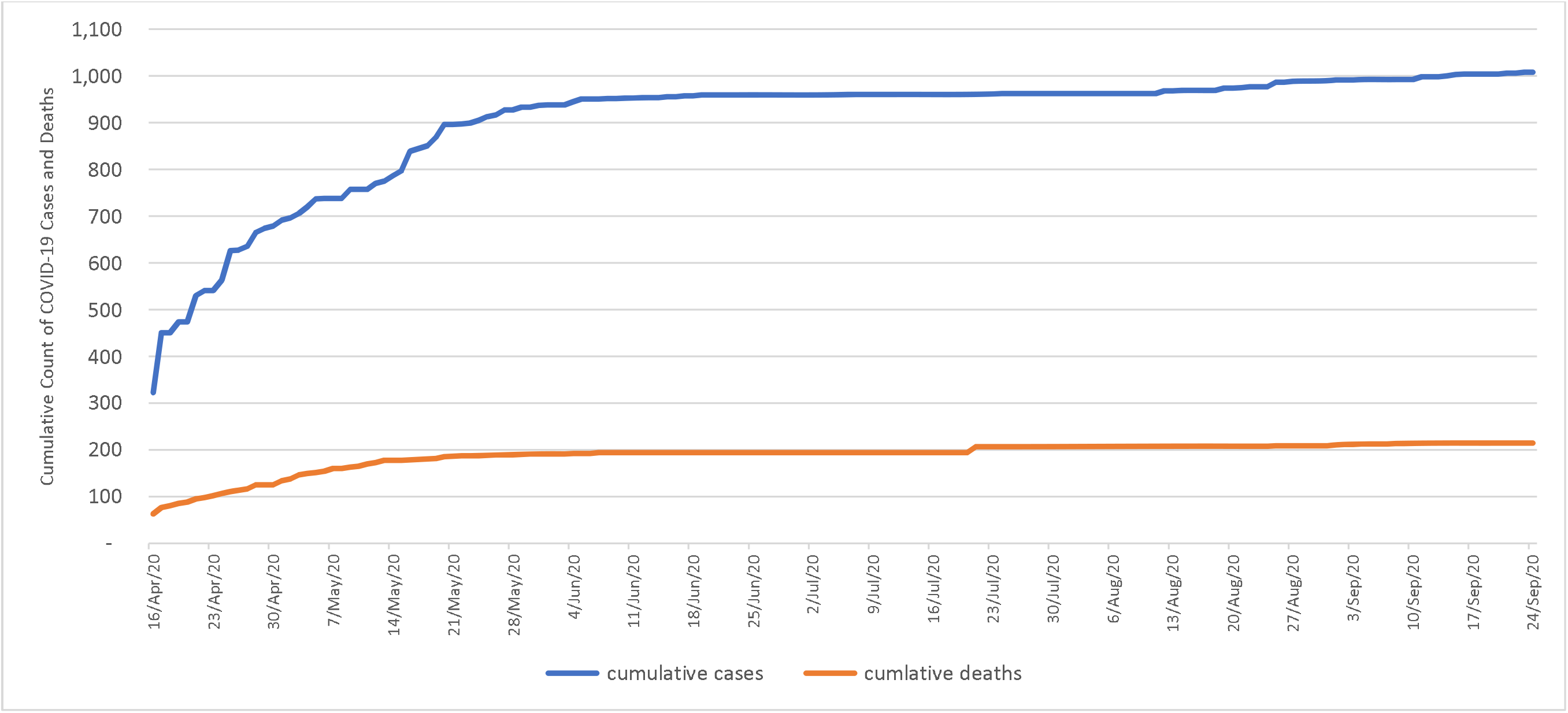
Cumulative Count of Retirement Home COVID-19 Cases and Deaths, Ontario, Mar 1-Sep. 24 2020 (N=770 facilities)

